# FOG2 coding variant Ser657Gly is associated with Coronary Microvascular Disease through altered hypoxia-mediated gene transcription

**DOI:** 10.1101/2023.05.22.23290352

**Authors:** M.A. Guerraty, Shefali Verma, Y.A. Ko, M.A. McQuillan, D. Conlon, J.W. Tobias, M.G. Levin, W. Haury, C. Zhang, R. Judy, Regeneron Genomics Center, PennMedicine Biobank, S. Tishkoff, S.M. Damrauer, Z. Arany, D.J. Rader

## Abstract

**Rationale:** The coronary microvasculature is crucial for proper cardiac function, and coronary microvascular disease (CMVD) has emerged as an underdiagnosed and undertreated cause of ischemic heart disease. Friend of GATA 2 (FOG2) is a transcriptional co-regulator crucial for coronary development and the maintenance of the coronary microvasculature in adult mice.Little is known about the role of FOG2 in humans or its role in CMVD.

**Objective:** Here, we report a genotype-first approach to determine the role of FOG2 in human coronary microvascular disease.

**Findings:** We performed Phenome-Wide association studies and deep cardiac phenotyping through the Electronic Health record in patients with FOG2 coding variants and identified an association between rs28374544 (A1969G, S657G) and CMVD. Patients with S657G had increased chest pain, smaller burden of obstructive coronary artery disease, and altered coronary blood flow. Differential gene and pathway analysis using several genomic datasets showed that carriers of S657G have increased expression of genes involved in angiogenesis, glycolysis, and the hypoxia-inducible factor (HIF) pathway. In vitro functional studies show that FOG2 S657G promotes angiogenic gene expression and angiogenesis while decreasing oxygen consumption rate. FOG2 also regulates HIF1a occupancy of angiogenic genes.

**Conclusions:** We identified a human missense variant which is associated with CMVD and established a potential mechanism by which the variant may altered angiogenic gene expression.

## Introduction

The coronary microvasculature regulates blood flow to meet cardiac metabolic demand and plays an important role in cardiac health and disease (1, 2). Coronary microvascular disease (CMVD), defined as disease of the coronary pre-arterioles, arterioles, and capillaries, can cause angina and myocardial infarction (1, 2). It also portends worse prognosis when present with coronary artery disease (CAD) (3) and has been implicated in the pathogenesis of heart failure with preserved ejection fraction (HFpEF) (4–6). Estimates suggest CMVD may account for 30-60% of ischemic heart disease (IHD) (7). Yet despite its clinical significance, little is known about the molecular pathways involved in CMVD.

Friend of GATA 2 (FOG2, also called zinc-finger protein motif 2) is a transcriptional coregulator which is crucial for coronary development (8). FOG2 has been shown to be required for the maintenance of coronary microvasculature in adult mice (9). In particular, in adult mice, FOG2 works in concert with GATA4 to promote expression of angiogenic genes (VEGFA, FGFs) and inhibit inhibitors of angiogenesis (TIMPs) (9). Some studies have linked FOG2 to congenital heart disease (10), however, little is known about the role of FOG2 in maintaining the coronary microvasculature in humans. Here we aim to determine whether FOG2 regulates the coronary microvasculature in humans and to test the hypothesis that dysregulation of FOG2 may contribute to CMVD.

Using a genotype-first approach focused on coding variants in FOG2 in participants in the Penn Medicine Biobank, we found that FOG2 S657G (rs28374544) was significantly associated with CMVD. We then used RNAseq datasets from human heart tissue and iPSCs lines to show that the variant is gain of function for angiogenic gene expression and identify potential pathways differentially regulated by the variant. Finally, we used *in vitro* studies to support a role for the variant in regulation of angiogenesis and cardiomyocyte metabolism.

## Results

### PheWAS identifies association between FOG2 S657G and clinical diagnosis of atherosclerosis

To evaluate for an association between *FOG2* and CMVD, we interrogated a cohort of 11,451 participants in the Penn Medicine BioBank (PMBB) who had undergone whole exome sequencing. There were 39 non-synonymous variants in the coding sequence or 3’ untranslated region (UTR) of FOG2 that were adequately represented in PMBB with at least 20 carriers of the variant (Supplemental Table 1). Phenome-Wide Association studies (PheWAS) were performed for each variant. After correcting for multiple testing, we identified one variant, FOG2 S657G, that was associated with PheCode for Atherosclerosis (p < 0.0001, Figure 1A, B). There were 44 patients homozygous for the minor allele (GG) and 556 patients heterozygous for the minor allele (AG), which represents a Minor Allele Frequency (MAF) of 0.03 in PMBB. This is comparable to other large heterogenous databases (GMAF, EXAC), however this variant is more common in populations of African ancestry (Supplemental Table 2). Indeed, 41/44 participants who were homozygous for the minor allele self-identified as African-American. GG patients were younger, more likely to be female and of African ancestry and with increased rates of hypertension and diabetes (Table 1). We further examined allele frequencies at this variant in 180 whole genome sequences from ethnically diverse African populations (11), as well as in 929 whole genomes from the Human Genome Diversity Project (HGDP)(12). We find that globally, the minor allele at rs28374544 (G) is only present at appreciable frequency within Africa (Figure 1E). Specifically, the variant is at high frequency in some populations from West Africa (Mandenka MAF = 0.32; Yoruba MAF = 0.25; Tikari MAF = 0.23), as well as in the Khoesan hunter-gatherers from Botswana (!Xoo MAF = 0.47; Ju|’Hoansi MAF = 0.37) and the Herero from Botswana (Herero MAF = 0.37). This allele is absent or at low frequency in other African regions (Figure 1F).

**Figure 1.**
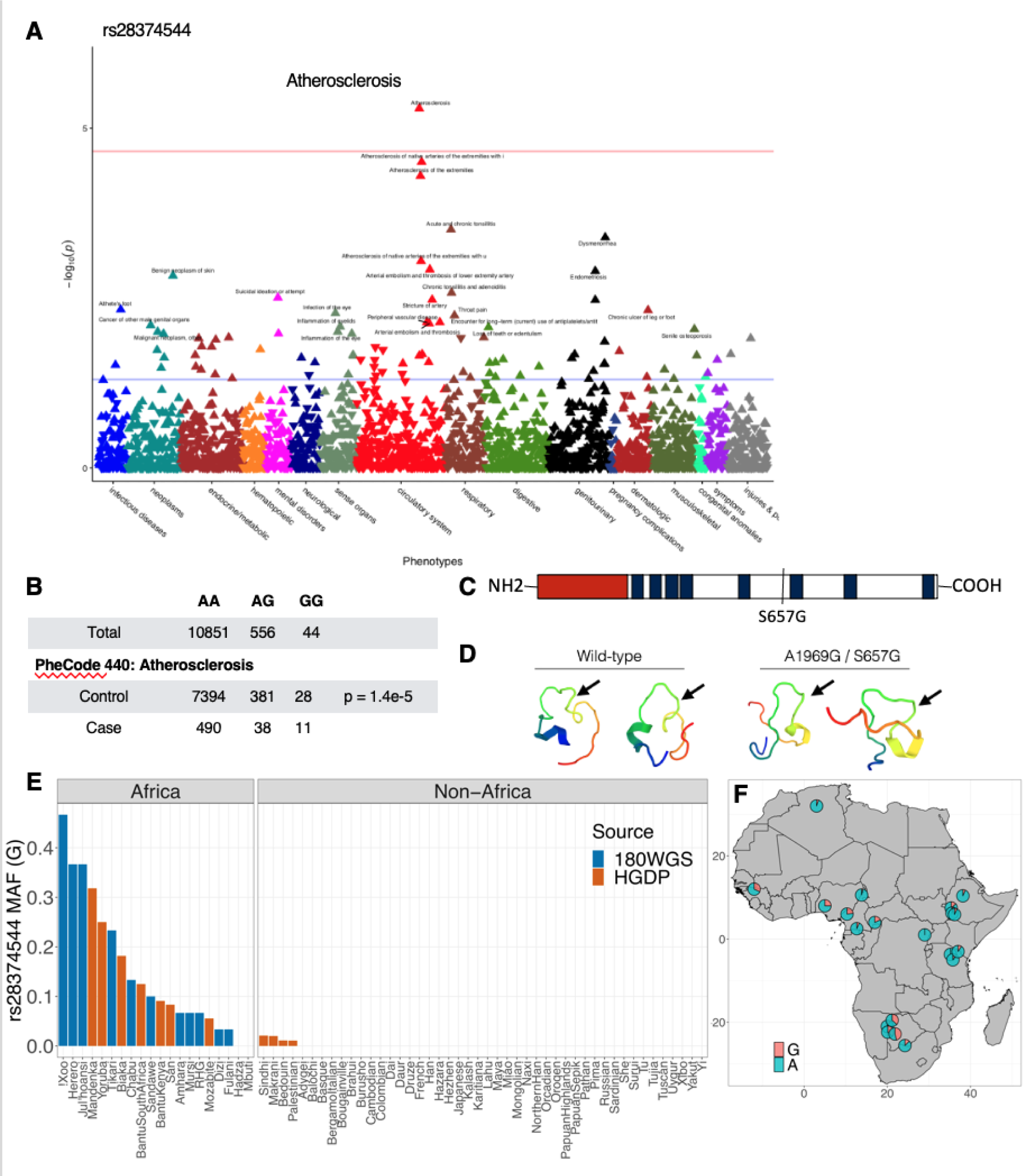
PheWAS identified an association between rs28374544 and a coronary vascular phenotype. (A) PheWAS analysis identified an association between rs28374544 and the Phecode for atherosclerosis. (B) Summary of the cases and controls of PheCode 440 in cases and controls. (C) Schematic of FOG2 protein highlighting the location of the coding variant relative zinc finger domains (blue) and repressor domain (red). (D) PEP-FOLD3 modeling showing local ring structure which is disrupted with S657G substitution. (E) MAF for the variant in African and Non-African populations. (F) Geographic distribution of the minor allele in Africa.

**Table 1.** PMBB demographics by genotype

|  | **Overall**<br>**(N = 11017)** | **AA**<br>**(N = 10436)** | **AG**<br>**(N = 537)** | **GG**<br>**(N = 44)** | **p-value<sup>2</sup>** |
| --- | --- | --- | --- | --- | --- |
| **Age** | 70 (61, 79) | 71 (62, 79) | 64 (55, 72) | 67 (57, 77) | <0.001 |
| **Sex** |  |  |  |  | <0.001 |
| F | 4,523 (41%) | 4,231 (41%) | 265 (49%) | 27 (61%) |  |
| M | 6,494 (59%) | 6,205 (59%) | 272 (51%) | 17 (39%) |  |
| **Race** |  |  |  |  |  |
| AM IND AK NATIVE | 1 (<0.1%) | 1 (<0.1%) | 0 (0%) | 0 (0%) |  |
| ASIAN | 139 (1.3%) | 139 (1.3%) | 0 (0%) | 0 (0%) |  |
| BLACK | 2,098 (19%) | 1,558 (15%) | 498 (93%) | 42 (95%) |  |
| HI PAC ISLAND | 5 (<0.1%) | 5 (<0.1%) | 0 (0%) | 0 (0%) |  |
| OTHER | 364 (3.3%) | 356 (3.4%) | 8 (1.5%) | 0 (0%) |  |
| UNKNOWN | 404 (3.7%) | 395 (3.8%) | 9 (1.7%) | 0 (0%) |  |
| WHITE | 8,006 (73%) | 7,982 (76%) | 22 (4.1%) | 2 (4.5%) |  |
| **Coronary atherosclerosis** | 4,360 (48%) | 4,174 (48%) | 167 (40%) | 19 (49%) | 0.008 |
| **Diabetes** | 3,174 (34%) | 2,915 (32%) | 236 (52%) | 23 (56%) | <0.001 |
| **Heart Failure** | 3,752 (42%) | 3,542 (42%) | 197 (43%) | 13 (37%) | 0.7 |
| **Hyperlipidemia** | 5,930 (64%) | 5,606 (64%) | 298 (64%) | 26 (65%) | >0.9 |
| **Hypertension** | 6,442 (69%) | 6,031 (69%) | 378 (79%) | 33 (80%) | <0.001 |
| **Ischemic heart disease** | 4,883 (51%) | 4,645 (51%) | 214 (46%) | 24 (55%) | 0.2 |
| ¹Median (IQR); n (%) |  |  |  |  |  |

²Kruskal-Wallis rank sum test; Pearson's Chi-squared test; Fisher's exact test

FOG2 is highly conserved between species, and rs28374544 represents missense variant A1969G, resulting in Ser657Gly substitution. This may introduce a flexible residue into an otherwise hydrophilic ring, potentially altering local protein structure. Using PEP-FOLD3 (13), we modeled 18 peptides flanking the variant and found that a glycine substitution introduces additional mobility into a loop domain between zinc fingers (Figure 1D).

Prior studies have reported an association between rs28374544 and complex congenital heart (CHD) disease (10). We used diagnoses codes and natural language processing from coronary angiography and echocardiography reports to determine whether participants who were heterozygous or homozygous for the minor allele had a history of complex congenital heart disease. We found that one participant heterozygous for the variant and two of the 44 participants homozygous for the variant had a history of Tetralogy of Fallot or Transposition of the Great Arteries and had undergone surgical repair in childhood. This further supports an association between CHD and rs28374544 with 4.5% of GG having CHD, (X^2^=31.2, p<0.0001, Supplemental Table 3).

### Clinical phenotypes of participants with FOG2 S657G

To gain insight into the association between genotype and the PheWAS association with atherosclerosis, we used the Electronic Health Record (EHR) to determine clinical cardiac phenotypes. First, for participants with prior clinical echocardiograms, we examined discrete echocardiographic parameters and found no association between genotype and left ventricular (LV) size, wall thickness or ejection fraction (Figure 2A,B). Similarly, there was no association between discrete hemodynamic parameters obtained on left heart catheterization including aortic blood pressure, heart rate, or LV filling pressures (Figure 2C). Despite this, participants with GG genotype were more likely to have inpatient admissions with primary diagnosis of chest pain (Figure 2D) or myocardial infarction (Figure 2E).

**Figure 2.**
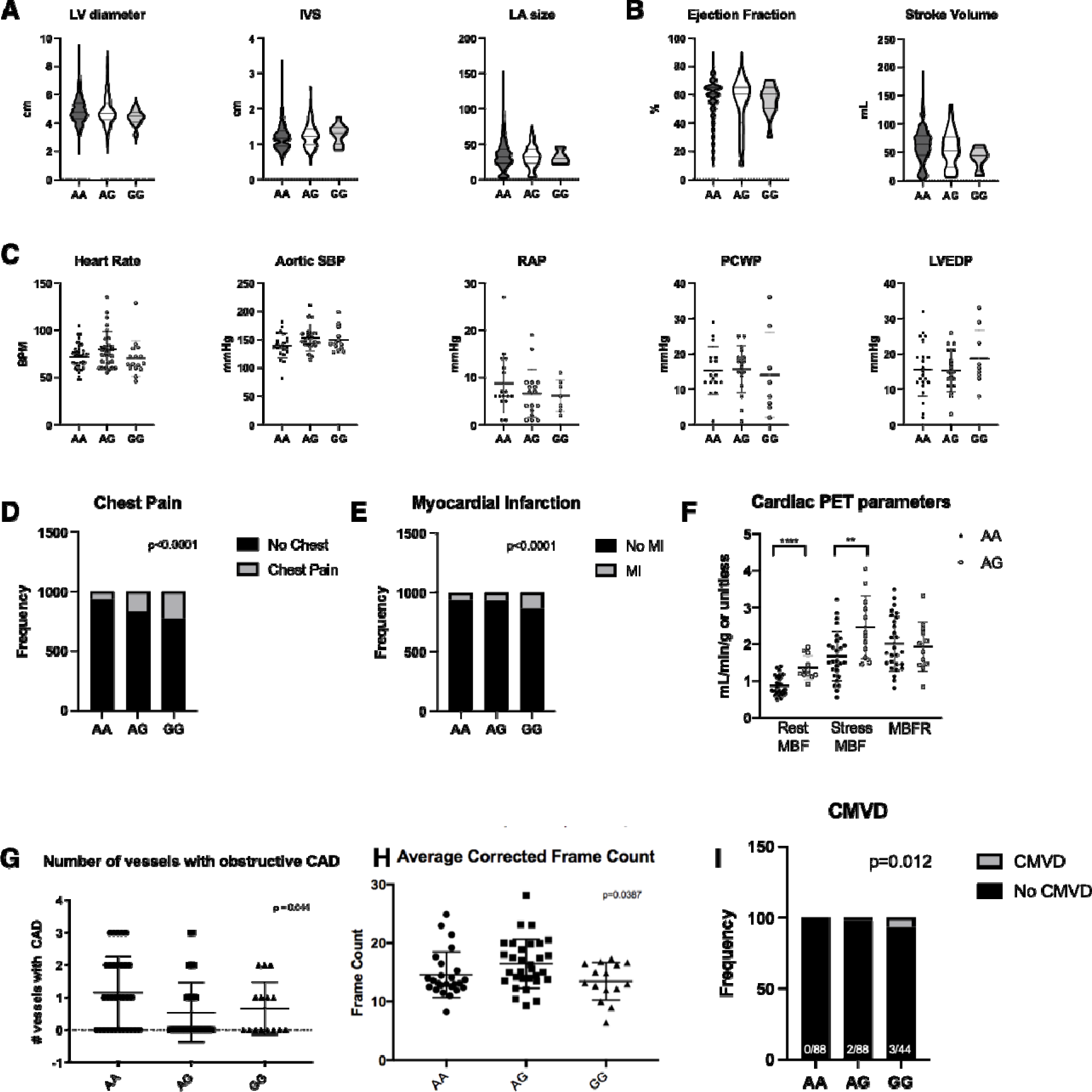
Clinical phenotyping of patients with and without FOG2 S657G variant. (A,B) Cardiac structure (left ventricular diameter and wall thickness and left atrial volume) and function (ejection fraction and stroke volume) assessed by echocardiography (n = 4748, 269, and 29 for AA, AG, GG, respectively). (C) Invasive hemodynamic parameters assessed during cardiac catheterization including heart rate aortic systolic blood pressure, right atrial pressure, pulmonary capillary wedge pressure, and LV end-diastolic pressure. (D, E) Frequency of inpatient primary diagnosis of chest pain and myocardial infraction. (F) Perfusion cardiac PET-measured quantitative flow. (G) Number of vessels with obstructive CAD in named coronary artery or first order branch by genotype. (H) Average corrected TIMI frame count by genotype. (I) Number of patients meeting clinical criteria for CMVD by genotype. (A-C, G, H, ANOVA analyses; D,E,I Chi-squared test, *p<0.005, **p<0.01).

Cardiac stress testing with Rubidium-82 perfusion Positron Emission Tomography (PET) imaging is routinely performed at the Hospital of University of Pennsylvania for clinical indications including the evaluation of chest pain. In addition to providing myocardial perfusion imaging, Rb-82 perfusion PET imaging provides quantitative myocardial blood flow (MBF) measurements at rest and under hyperemic, or cardiac stress, conditions. The ratio of stress MBF to rest MBF is the MBF reserve (MBFR) which, in the absence of coronary stenosis, reflects coronary microvascular function. Since self-identified race is known to affect MBFR (14), we examined MBF values for African-American participants with AA and AG genotype in the PMBB. We found that patients heterozygous for the variant had increased ∼30% increased resting MBF (mean 1.02 vs 1.36 ml/min/g, 95% CI [0.11, 0.57], p<0.005) and increased stress MBF (mean 1.85 vs 2.47 ml/min/g, 95% CI [0.17, 1.07], p<0.01), with no change in MBFR (Figure 2F). This suggests a genotype-specific effect on myocardial blood flow.

### Cardiac phenotyping of matched cohorts reveal association between FOG2 S657G and CMVD

To perform more in-depth cardiac phenotyping, cohorts of participants with AA and AG genotype were matched to GG participants based on age, gender, and self-reported race (Table 2). The groups had similar distribution of cardiovascular risk factors and differed only in diagnosis of atherosclerosis, which is consistent with the PheWAS analysis.

**Table 2.** Cohorts matched on age, sex and ancestry.

|  | **AA**<br>**(N = 88)** | **AG**<br>**(N = 88)** | **GG**<br>**(N = 44)** |
| --- | --- | --- | --- |
| Age | 58.3 (15.0) | 58.0 (15.0) | 58.0 (15.2) |
| Female | 54 (61.4) | 54 (61.4) | 27 (61.4) |
| African-American | 82 (93.2) | 82 (93.2) | 41 (93.2) |
| Body Mass Index (kg/m<sup>2</sup>) | 32.7 (8.6) | 31.3 (7.2) | 32.9 (9.5) |
| Hypertension | 76 (86.4) | 71 (80.7) | 35 (79.5) |
| History of Myocardial Infarction | 2 (2.3) | 4 (4.5) | 1 (2.3) |
| History of Prior CAD | 37 (42.0) | 38 (43.2) | 22 (50) |
| Diabetes | 0 (0) | 0 (0) | 0 (0) |
| Chronic Kidney Disease | 8 (9.1) | 8 (9.1) | 3 (6.8) |
| <sup>†</sup> Median (IQR); n (%) |  |  |  |

The burden of CAD was assessed based on manually abstracted coronary angiograms by a blinded reviewer. Obstructive CAD was defined as ≥70% stenosis in a named coronary artery or first-order branch or >50% stenosis in the left main coronary artery. GG participants had decreased burden of CAD (Figure 2G). Thrombolysis in Myocardial infarction (TIMI) Frame Count (TFC) is a quantitative measure of how long it takes contrast dye to travel from a proximal landmark to a distal landmark during a coronary angiogram cine and has been previously used as a measure of the coronary microvasculature (3, 15). We found that participants with the reference allele had TFC of 14.6 ± 3.88 whereas those heterozygous or homozygous for the variant had TFC of 16.46 ± 4.17 and 13.47 ± 3.20, respectively (ANOVA p=0.039, Figure 2H). A clinical diagnosis of CMVD is based on three features: (1) symptoms of cardiac ischemia including chest pain, angina, or shortness of breath, (2) no other obvious cardiac cause of symptoms, such as valvular disease or cardiomyopathy, and (3) no obstructive coronary artery disease. We assessed participants for clinical criteria of CMVD and found that 6.8% of those homozygous for the variant met CMVD criteria whereas 2.3% and 0% of those heterozygous for or without the variant met criteria (Figure 2I).

### Genomic analysis of FOG2 S657G in human heart tissue and induced Pluripotent Stem Cell Lines

We next sought to assess the effects of FOG2 S657G on gene expression in human heart tissue. We used RNAseq and genotyping data from control hearts from African-American patients from the MAGnet study (Figure 3A)(16). We further subset by sex and focused on females since there were more female samples and a majority of our human cohort was female. First, we established the effect of FOG2 S657G on known FOG2 target genes. Unbiased clustering of genes from Zhou et al (9) showed that angiogenic and anti-angiogenic gene targets of FOG2 clustered separately, with the exception of *FGF16* and *COL4A3* (Figure 3B). AG and GG genotypes were associated with increased expression of angiogenic genes including *VEGFA*, *FGF2*, *FGF9* and decreased expression of inhibitors of angiogenesis such as *TIMP1* and *TIMP2*. These data suggest that FOG2 S657G may act as a gain of function variant for angiogenic genes. We next performed differential gene expression analysis comparing AA females with AG and GG females. Gene set enrichment analysis of differentially expressed genes identified several pathways as differentially regulated in patients with rs283, including unfolded protein response, hypoxia, and MTORC1 pathways (Figure 3C).

**Figure 3.**
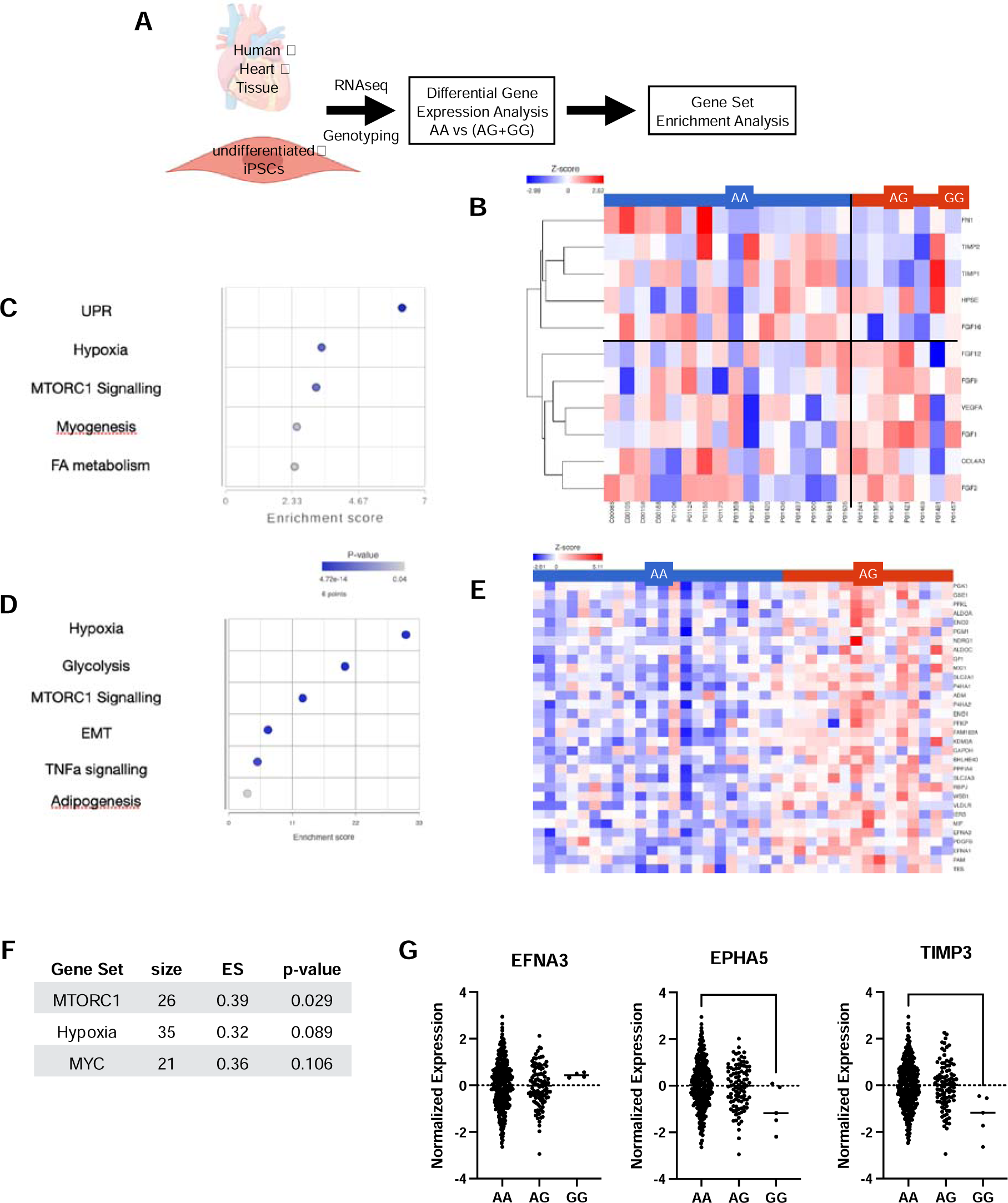
Differential gene and protein expression and pathway analysis in human heart tissue, undifferentiated iPSC, and plasma from individuals with and without the variant. (A) Differential gene expression analysis and gene set enrichment analysis was performed using RNAseq expression data from previously published human heart tissue and undifferentiated iPSCs datasets. (B) Heatmap from human heart tissue gene expression in patients with the variant (AA, blue bar) and (AG or GG, red bar) for known gene targets of FOG2. Angi-angiogenic genes cluster together and are decreased in individuals with the variant (top left quadrant) whereas angiogenic genes cluster together and are generally increased (bottom right quadrant). (C, D) Gene Set Enrichment Analysis using hallmark gene sets identified Hypoxia and MTORC1 signaling as top differentially regulated sets. (E) Heatmap of genes from the hypoxia gene sets in undifferentiated iPSC cells highlights increased expression of regulators of glycolysis including *ALDOA*, *ALDOC*, *PFKL*, *PGK1*, and *ENO2. EFNA3* was also increased in this gene set. (F) Top gene sets identified using a previously reported plasma protein data set also identified MTORC1 with nominal p-value 0.029. (G) Normalized protein expression for angiogenic proteins EFNA3 suggests a trend towards increase. EPHA5 and TIMP3 were decreased (ANOVA with multiple comparison testing, *p<0.05, **p<0.01)

To confirm these results and determine whether these changes represented fundamental cell-autonomous processes or were the result of interactions of multiple cell-types, we assessed the effects of FOG2 S657G on gene expression in a cohort of previously reported undifferentiated human iPSC lines. We analyzed the RNAseq data from 25 cell lines isolated from African American female participants: 9 lines from carriers homozygous for the variant and 16 lines from individuals without the variant. Gene-set enrichment analysis identified hypoxia, glycolysis and MTROC1 pathways as the most significantly enriched gene sets (Figure 3D). Several of the hypoxia related genes are increased in AG which suggests that FOG2 may contribute to the regulation of hypoxia-inducible genes. Several angiogenic and hypoxia-inducible gene targets were increased in AG females, including *ADM*, *EFNA1*, *EFNA3* and *PDGFB*. Several hypoxia-regulated genes which play a role in metabolism were also increased in AG females, including *ALDOA*, *ALDOC*, *PFKL*, *PGK1*, and *ENO2*.

Lastly, we examined the levels of serum proteins in 389 PMBB participants of African ancestry who had undergone proteomic profiling with SOMAscan Somalogic and genotyping. Mean age of participants was 61+/-13.6, and 58% were male. For this exploratory analysis, we compared the top plasma proteins differentially regulated between AA and AG or GG, and performed gene set enrichment analysis. MTORC1 was the most enriched pathway, and hypoxia was second, though not statistically significant (Figure 3F). EFNA3 which was increased in iPSCs showed a trend towards increase in GG participants. EPHA5, a receptor for EFNA3 was downregulated in participants homozygous for the variant. TIMP3, an anti-angiogenic gene, was decreased in the serum of GG participants (Figure 3G).

### FOG2 S657G regulates hypoxia-induced gene expression

To determine whether the effects of FOG2 S657G variant may be due to the variant alone rather than other variants which may be in linkage disequilibrium, we turned to an *in vitro* system. AC16 cardiomyocytes, an immortalized human left ventricular cardiomyocyte cell line (17), underwent lipid-based transfection to overexpress control empty vector, FOG2, or FOG2 S657G. We saw no significant differences under basal conditions. Based on genomic data suggesting FOG2 may be involved in hypoxia-related pathways, we treated cells with low dose dimethyloxallyl glycine (DMOG), a chemical stabilizer of HIF proteins. We then noted an increase in angiogenic genes *VEGFA* and *ADM* in cells treated with FOG2 S657G relative to wild type FOG2. *ANGPT1*, which is known to decrease angiogenesis and promote vessel maturation, was decreased by FOG2 S657G (Figure 4A). To assess the functional effect of these gene changes on angiogenesis, we used conditioned media from the AC16 cells in a tube formation assay. Conditioned media from cells treated with FOG2 S657G formed tube networks with more junctions and segments and longer branches and total length (Figure 4B,C), consistent with increased angiogenic potential.

**Figure 4.**
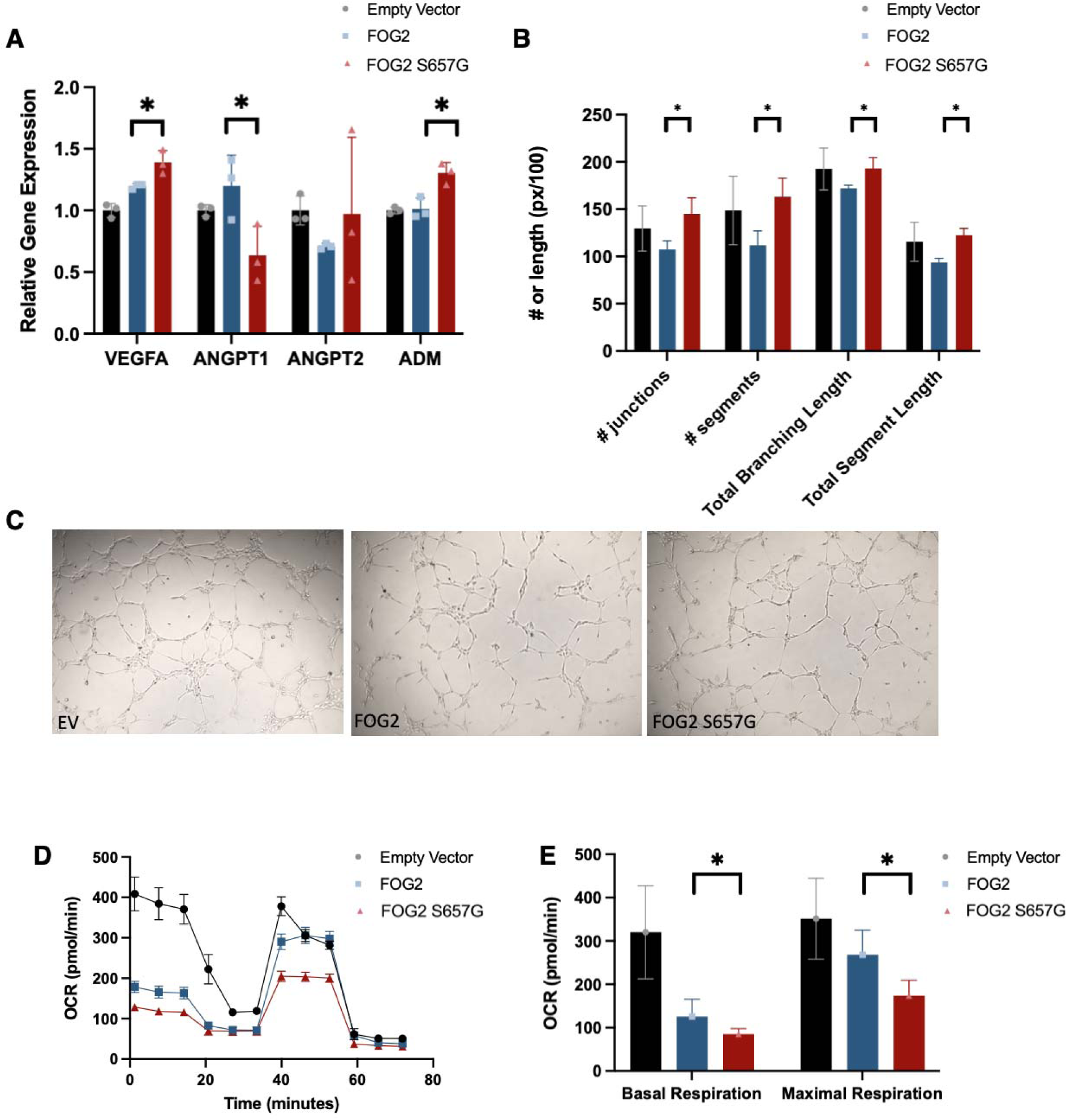
FOG2 S657G regulates angiogenesis and metabolism *in vitro*. (A) Angiogenic gene expression in AC16 cardiomyocytes overexpressing FOG2 S657G relative to wild-type FOG2. (B,C) Tube formation angiogenesis assay using conditioned media from cells treated with FOG2 S657G, wild-type FOG2, or control empty vector. (D, E) Oxygen consumption rate measured during a Mito Stress Test in AC16 cells. ANOVA analyses with multiple comparison testing, *P<0.5)

Genomic data suggested that FOG2 S657G promoted expression of glycolytic genes. To determine whether FOG2 S657G had an effect on oxidative phosphorylation, we measured oxygen consumption rate (OCR). FOG2 S657G-overexpressing cells had decreased basal respiration. Following FCCP administration to measure maximal oxygen respiration, FOG2 S657G-overexpressing cells had lower OCR.

### FOG2 regulates expression of angiogenic HIF1a targets

Genomic data suggested a link between FOG2 S657G and hypoxia-regulated pathways, and *in vitro* studies suggested FOG2 S657G may regulate HIF-induced angiogenesis and oxidative phosphorylation. Though FOG2 is known to interact with GATA4 (18) and additional transcription factors including COUP-TF2 (19), it is now known whether FOG2 can regulate HIF1a-mediated gene transcription. We used CRISPR-Cas9 gene editing to generate AC16 cardiomyocyte cell lines deficient in FOG2. We targeted exon 8 which resulted in decrease of both FOG2 and isoform FOG2S (Figure 5A). We treated cells with DMOG to induce HIF1a-mediated gene expression. Cells deficient in FOG2 showed a decrease in *VEGFA* and *ADM* transcripts (Figure 5B). In contrast to angiogenic HIF1a targets, metabolic gene targets *PDK1* and *BNIP3* were similar in control and FOG2 knockdown cell lines (Figure 5C). We next performed cleavage under targets and release using nuclease (CUT&RUN) to profile HIF1a-chromatin interactions in the promoter region of these genes. There was a region of VEGFA and ADM promoters with a focal decrease in HIF1a binding in FOG2KD cell lines. In contrast, HIF1a occupancy in the promoters of PDK1 and BNIP3 was similar in control and FOG2 KD cells. These data suggest that FOG2 may serve as an angiogenic-gene-specific coregulator of HIF1a.

**Figure 5.**
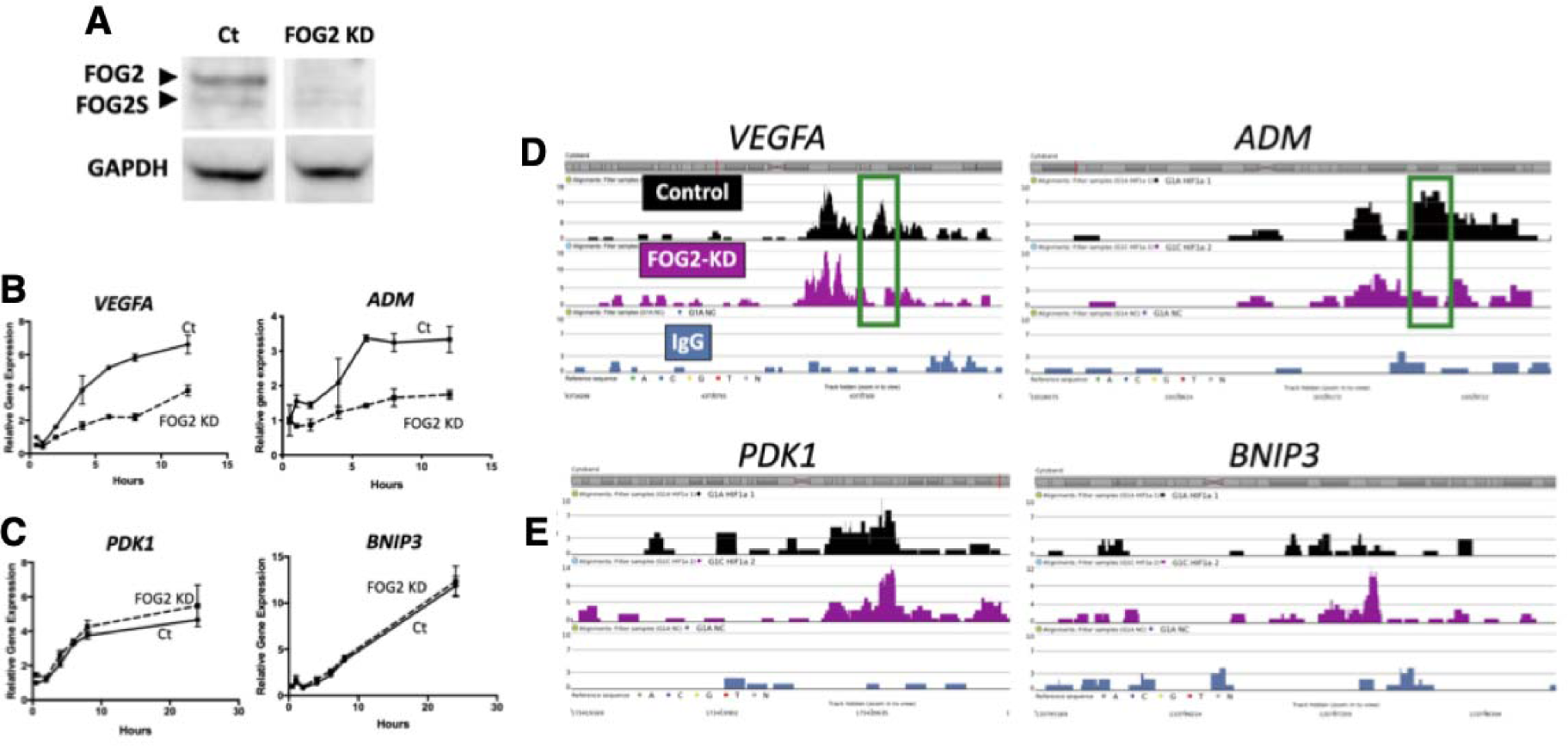
FOG2 knockdown (KD) blunts hypoxia-induced angiogenic gene expression. (A) Western Blot showing decreased FOG2 in FOG2-KD cell line, generated using used CRISPR-Cas-mediated disruption of exon 8 which is shared by FOG2 and FOG2S. (B) When cells are exposed to hypoxia, FOG2 KD show decreased expression of angiogenic genes *VEGFA* and *ADM* over time relative to control. (C) Gene expression of *PDK1* and *BNIP3*, non-angiogenic targets of HIF1a, is not affected in FOG2-KD. (D, E) CUT&RUN and DNA sequencing shows HIF1a binding to the promoters of *VEGFA*, *ADM*, *PDK1*, and *BNIP3*. FOD2KD had decreased occupancy relative to control (green boxes).

## Discussion

We use a genotype-first approach to show an association between a coding variant FOG2 S657G in the FOG2 gene and clinical presentations and imaging findings consistent with CMVD. Using gene expression data from human heart samples, we show that FOG2 S657G is a gain of function variant which is associated with altered angiogenic gene expression. Unbiased analyses in both human heart tissue and iPSC cells suggest that FOG2 S657G is associated with gene changes related to hypoxia, metabolism, and unfolded protein response pathways. *In vitro* functional studies support a role for FOG2 S657G regulation of hypoxia-induced angiogenic gene expression, tube formation, and cellular metabolism.

Based on heart tissue gene expression, FOG2 S657G may serve as a gain of function variant to promote angiogenesis. However, the mechanism by which FOG2 S657G may lead to CMVD is unknown. One potential explanation may be that increased angiogenic growth factors alter vessel formation resulting in inadequate coronary microvasculature. For example, Pontes-Qureo et al shown that increased angiogenic signals can lead to dysregulation of angiogenesis (20). Another possibility is that FOG2 S657G impairs cardiac metabolism and disrupts the tight coupling blood flow and cardiomyocyte metabolism. Any mismatch between blood supply and cardiac demand can result in cardiac ischemia and symptoms of CMVD. Most likely, however, both contribute to the phenotype. Additional mechanistic studies in *in vivo* models are needed to untangle the relative contributions of these on coronary microvasculature and *in vivo* cardiac metabolism.

The mechanism by which FOG2 regulates gene expression is not clear. On one hand, FOG2 is known to interact with GATA4, and the S657G variant has been shown to reduce the ability of FOG2 to repress GATA4-mediated transcription (10). As such, FOG2 serves as a transcriptional co-repressor, possible via recruitment of NURD complex (21). Conversely, Zhou et al showed that FOG2 interaction with GATA 4 is necessary for regulation of angiogenic FOG2 targets such that disrupting GATA4-FOG2 interactions decreased target gene expression. Thus FOG2 regulation must be multifactorial and likely involve interactions with other cardiac transcription factors. In our studies, we found that many hypoxia-inducible genes are regulated by FOG2. We also show that a FOG2-knockout cell line has a blunted response to hypoxia-mimicking stimulus and leads to decreased HIF1a occupancy in target gene promoters. These data suggest FOG2 may serve as a cofactor for hypoxia-inducible factor 1a. Additional studies are needed to understand these interactions.

FOG2 S657G is also unique in that it is highly enriched, if not exclusively present, in populations of African ancestry. It is well established that African American patients suffer from a higher burden of cardiovascular disease and worse cardiovascular outcomes (22). These differences are largely driven by social determinants of health. It is also possible that underlying genetic variation interacts with environmental factors to promote CMVD. Additionally, CMVD has been implicated in the pathogenesis of Heart Failure with Preserved Ejection fraction (HFpEF). Patients with FOG2 S657G may therefore be predisposed to not only angina and myocardial infarction, but also HFpEF. Understanding the mechanisms driving these differences can help identify novel solutions and chip away at health disparities.

There are several limitations to this work. It is a single center study performed at a tertiary care center. Additionally, though our results suggest a link with human phenotype and functional studies support a role for FOG2 S657G in mediating hypoxia-related pathways, it is also possible that FOG2 S657G is in linkage disequilibrium with other variants that regulate FOG2 expression or protein function direction or regulate expression of other genes. We also do not directly show that cardiac expression of FOG2 S657G is driving the phenotype seen in patients with the variant. FOG2 is expressed in few tissues, including the heart, liver, cerebellum, and ovaries in the Genotype-Tissue Expression project (23). Our *in vitro* studies outline a plausible mechanism by which FOG2 may affect cardiomyocyte gene expression and result in a CMVD-like phenotype. However, *in vivo* studies are needed to definitively establish the role of FOG2 S657G on coronary microvasculature and fully understand the interplay between cardiac cell types. Lastly, we outline a potential role whereby FOG2 S657G may lead to dysregulation of angiogenic factors which may affect coronary microvasculature. This could affect coronary microvascular structure and function in adults, but it is also possible that the variant results in altered coronary patterning during development. Additional studies are needed to understand whether FOG2 S657G may affect coronary microvascular development.

In summary, we examined FOG2 variants in the Penn Medicine Biobank and identified a coding variant in FOG2 gene which is predominantly present in patients of African descent and is associated with a CMVD-like phenotype. These results establish an association between FOG2 gene and CMVD and suggest a potential mechanism whereby FOG2 may regulate HIF target gene expression. Future studies are needed to further characterize potential interactions between FOG2 and HIFs and to understand the role of altered angiogenic gene expression on coronary microvascular structure and function.

## Methods

### Study participants

The Penn Medicine BioBank (PMBB) included more than 90,000 participants from the University of Pennsylvania Health System (24). Appropriate consent was obtained from each participant regarding storage of biological specimens, genetic sequencing, and access to all available electronic health record (EHR) data. The study was approved by the Institutional Review Board of the University of Pennsylvania and complied with the principles set out in the Declaration of Helsinki.

### PheWAS

We used logistic regression to identify phenotypic associations for the genetic variant of interest, using PheCodes as the dependent variable, and age, age-squared, sex, and the first 10 PCs from genotypes as covariates. We selected phenotypes that have 5% prevalence, which corresponded to 20 cases per PheCode.

### Cardiac phenotyping

Discrete data from coronary angiograms and echocardiograms was obtained for patients who had undergone testing as part of routine clinical care. If more than one study was available, then the most complete and recent study was used. ICD9/10 codes for chest pain and myocardial infarction were used to identify patients presenting with these clinical syndromes. Perfusion PET stress testing was performed as part of routine medical care as previously described (14). For deeper phenotyping of matched cohorts, coronary angiogram data was manually abstracted by a blinded operator. Degree of coronary stenosis was assessed in all named coronary arteries and first order branches. Obstructive disease was defined as ≥ 50% stenosis in the left main coronary artery or ≥ 70% in a named coronary or first order branch. Thrombolysis in Myocardial Infarction (TIMI) frame count was performed by a blinded operator as previously described (3). The corrected frame count for all non-obstructed vessels was averaged to represent the frame count for a given patient.

### Gene expression and Gene Set Enrichment Analysis

Gene expression data was obtained from the Gene Expression Omnibus Repository for (16, 25). Partek® Flow® software (version 10.0.23.0414) was used for differential gene expression analysis (DESeq2), hierarchical clustering and heat map generation, and Gene Set Enrichment Analysis using the hallmark pathway gene set (26).

### Protein measurements

Plasma was extracted and stored at −80 °C before shipping on dry ice to SomaLogic, Inc. (Boulder, Colorado, US) for measurement of the relative concentration of 1,305 proteins using the multiplexed, aptamer-based SOMAscan assay. See Supplemental Methods.

### In vitro assays

AC16 cardiomyocytes (SCC109, Sigma Millipore, Burlington, MA) were transfected using Lipofectamine 3000 reagent (Invitrogen, Waltham, MA). Gene expression was assayed using qRT-PCR (Applied Biosciences). Conditioned media from transfected AC16 cardiomyocytes was used in a tube formation assay. Briefly, HUVECs were seeded onto growth-factor restricted Matrigel, cultured with conditioned media, and imaged at 4 hours. Oxygen Consumption Rate was measured using a Seahorse Bioanalyzer XF and Mito Stress Test reagents (Agilent, Santa Clara, CA). CRISPR-Cas9 was used to create a FOG2-deficient AC16 cardiomyocyte cell line. Cleavage Under Target and Reac Under Nucleosome (CUT&RUN) kit (CST #86652) was performed using with HIF1a (CST #36169S and Abcam #2185) or control IgG antibodies (CST #66362). Sequencing was performed on NovaSeq and data were analyzed using Partek Flow software (version 10.0.23.0414). See Supplemental Methods for additional details.

## Data Availability

All data produced in the present study are available upon reasonable request to the authors.

## Author Contributions

MG: experimental design, conducting experiments, data acquisition and analysis, and manuscript writing

SV: conducting experiments, data generation, analysis, and visualization, manuscript editing

YAK: conducting experiments, data generation, analysis, and visualization, manuscript editing

M.A.M: data analysis and visualization, manuscript editing

DC: conducting experiments, data acquisition and analysis, manuscript editing

MGL: data generation, analysis, and visualization, manuscript editing

Z.C.: data analysis and manuscript editing

WH: data generation, manuscript editing

RJ: data generation, manuscript editing

RGC: data generation, manuscript editing

ST, SMD, ZA, DJR: scientific input, manuscript editing

## Acknowledgements

We would like to acknowledge the contributions of Mike Morley for assistance with the MAGnet data, Susannah Elwyn for support with *in vitro* studies and sequencing, and the University of Pennsylvania Next Generation Sequencing Core. We acknowledge the Penn Medicine BioBank (PMBB) for providing data and thank the patient-participants of Penn Medicine who consented to participate in this research program. We would also like to thank the Penn Medicine BioBank team and Regeneron Genetics Center for providing genetic variant data for analysis. The PMBB is approved under IRB protocol# 813913 and supported by Perelman School of Medicine at University of Pennsylvania, a gift from the Smilow family, and the National Center for Advancing Translational Sciences of the National Institutes of Health under CTSA award number UL1TR001878.

