## Supplemental Files for "FOG2 coding variant Ser657Gly is associated with Coronary Microvascular Disease through altered hypoxia-mediated gene transcription"

**Supplemental Data**

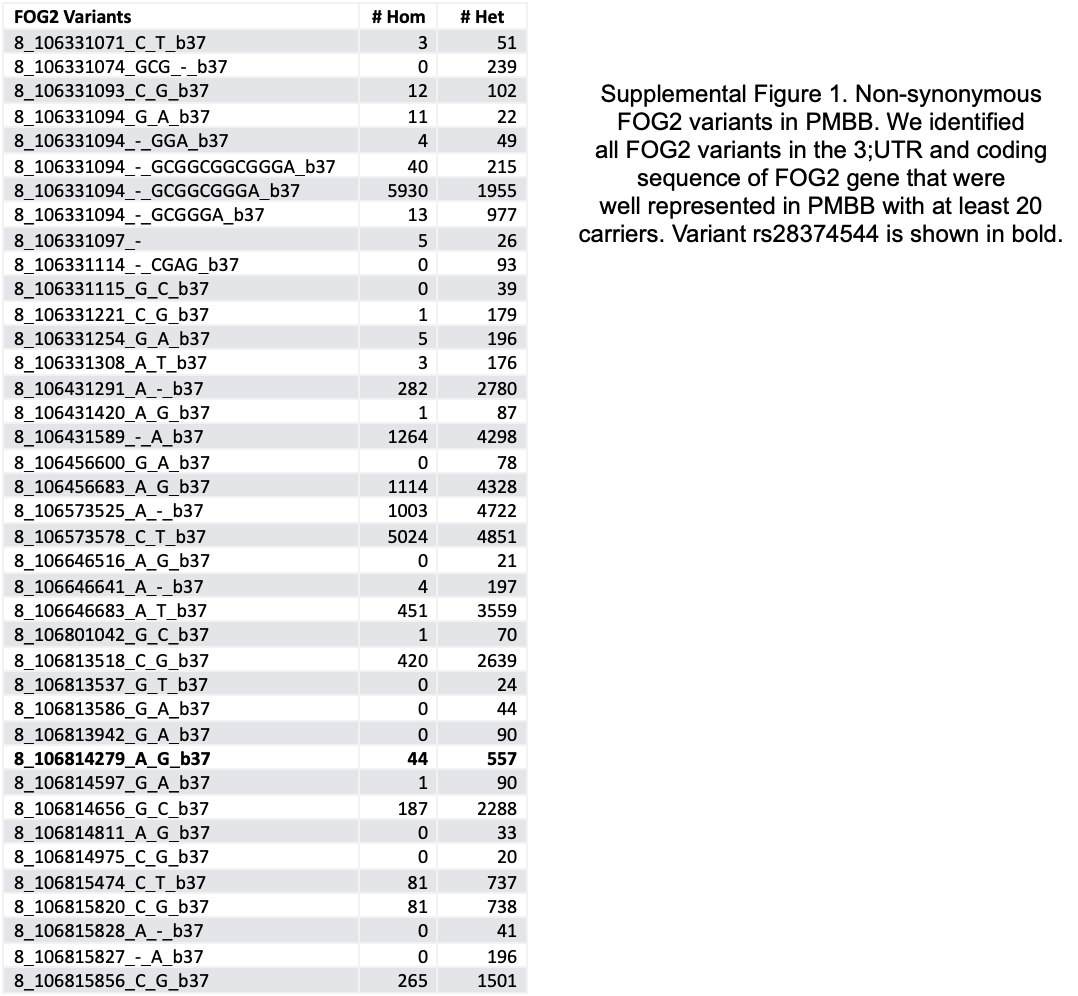

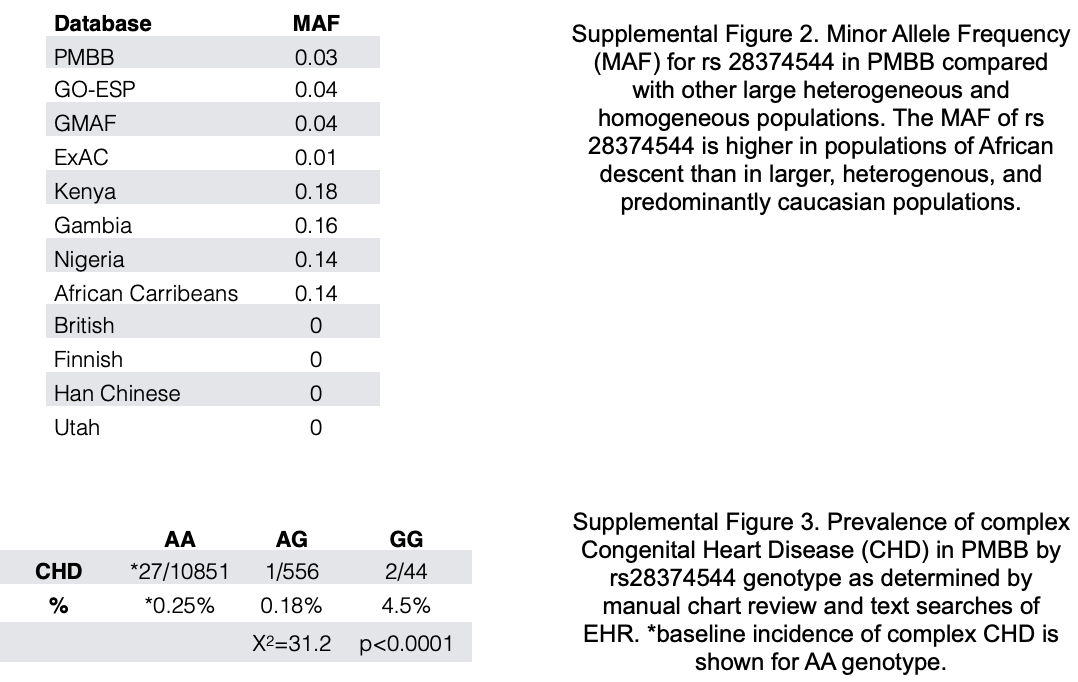

**Supplemental Note
Penn Medicine BioBank Banner Author List and Contribution Statements**

**PMBB Leadership Team**

Daniel J. Rader, M.D., Marylyn D. Ritchie, Ph.D.

Contribution: All authors contributed to securing funding, study design and oversight. All authors reviewed the final version of the manuscript.

**Patient Recruitment and Regulatory Oversight**

JoEllen Weaver, Nawar Naseer, Ph.D., M.P.H., Afiya Poindexter, Khadijah Hu-Sain, Yi-An Ko, Ph.D.

Contributions: JW manages patient recruitment and regulatory oversight of study. NN manages participant engagement, assists with regulatory oversight, and researcher access. AP, KH, YK perform recruitment and enrollment of study participants.

**Lab Operations**

JoEllen Weaver, Meghan Livingstone, Fred Vadivieso, Stephanie DerOhannessian, Teo Tran, Julia Stephanowski, Monica Zielinski, Ned Haubein, Joseph Dunn

Contribution: JW, ML, FV, SD conduct oversight of lab operations. ML, FV, AK, SD, TT, JS, MZ perform sample processing. NH, JD are responsible for sample tracking and the laboratory information management system.

**Clinical Informatics**

Anurag Verma, Ph.D., Colleen Morse Kripke, M.S. DPT, MSA, Marjorie Risman, M.S., Renae Judy, B.S.

Contribution: All authors contributed to the development and validation of clinical phenotypes used to identify study subjects and (when applicable) controls.

**Genome Informatics**

Anurag Verma Ph.D., Shefali S. Verma, Ph.D., Yuki Bradford, M.S., Scott Dudek, M.S., Theodore Drivas, M.D., Ph.D.

Contribution: A.V., S.S.V. are responsible for the analysis, design, and infrastructure needed to quality control genotype and exome data. Y.B. performs the analysis. T.D. and A.V. provides variant and gene annotations and their functional interpretation of variants.

Regeneron Genetics Center Banner Author List and Contribution Statements

**RGC Management and Leadership Team**

Goncalo Abecasis, PhD , Aris Baras, M.D. , Michael Cantor, M.D. , Giovanni Coppola, M.D. , Andrew Deubler , Aris Economides, Ph.D. , Luca A. Lotta, M.D., Ph.D. , John D. Overton,Ph.D., Jeffrey G. Reid, Ph.D. , Katherine Siminovitch, M.D. , Alan Shuldiner, M.D.

**Sequencing and Lab Operations**

Christina Beechert , Caitlin Forsythe, M.S. , Erin D. Fuller , Zhenhua Gu, M.S. , Michael Lattari , Alexander Lopez, M.S. , John D. Overton, Ph.D. , Maria Sotiropoulos Padilla, M.S. , Manasi Pradhan, M.S. , Kia Manoochehri, B.S. , Thomas D. Schleicher, M.S. , Louis Widom , Sarah E. Wolf, M.S. , Ricardo H. Ulloa, B.S.

**Clinical Informatics**

Amelia Averitt, Ph.D. , Nilanjana Banerjee, Ph.D. , Michael Cantor, M.D. , Dadong Li, Ph.D. , Sameer Malhotra, M.D. , Deepika Sharma, MHI , Jeffrey Staples , Ph.D.

**Genome Informatics**

Xiaodong Bai, Ph.D. , Suganthi Balasubramanian, Ph.D. , Suying Bao, Ph.D. , Boris Boutkov, Ph.D. , Siying Chen, Ph.D. , Gisu Eom, B.S. , Lukas Habegger, Ph.D. , Alicia Hawes, B.S. , Shareef Khalid , Olga Krasheninina, M.S. , Rouel Lanche, B.S. , Adam J. Mansfield, B.A. , Evan K. Maxwell, Ph.D. , George Mitra, B.A. , Mona Nafde, M.S. , Sean O’Keeffe, Ph.D. , Max Orelus, B.B.A. , Razvan Panea, Ph.D. , Tommy Polanco, B.A. , Ayesha Rasool, M.S. , Jeffrey G. Reid, Ph.D. , William Salerno, Ph.D. , Jeffrey C. Staples, Ph.D. , Kathie Sun, Ph.D.

**Analytical Genomics and Data Science**

Goncalo Abecasis, D.Phil. , Joshua Backman, Ph.D. , Amy Damask, Ph.D. , Lee Dobbyn, Ph.D. , Manuel Allen Revez Ferreira, Ph.D. , Arkopravo Ghosh, M.S. , Christopher Gillies, Ph.D. , Lauren Gurski, B.S. , Eric Jorgenson, Ph.D. , Hyun Min Kang, Ph.D. , Michael Kessler, Ph.D. , Jack Kosmicki, Ph.D. , Alexander Li , Ph.D. , Nan Lin, Ph.D. , Daren Liu, M.S. , Adam Locke, Ph.D. , Jonathan Marchini, Ph.D. , Anthony Marcketta, M.S. , Joelle Mbatchou, Ph.D. , Arden Moscati, Ph.D. , Charles Paulding, Ph.D. , Carlo Sidore, Ph.D. , Eli Stahl, Ph.D. , Kyoko Watanabe, Ph.D. , Bin Ye, Ph.D. , Blair Zhang, Ph.D. , Andrey Ziyatdinov, Ph.D.

**Therapeutic Area Genetics**

Ariane Ayer, B.S. , Aysegul Guvenek, Ph.D. , George Hindy, Ph.D. , Giovanni Coppola, M.D. , Jan Freudenberg, M.D. , Jonas Bovijn M.D. , Katherine Siminovitch, M.D. , Kavita Praveen, Ph.D. , Luca A. Lotta, M.D. , Manav Kapoor, Ph.D. , Mary Haas, Ph.D. , Moeen Riaz , Ph.D. , Niek Verweij, Ph.D. , Olukayode Sosina, Ph.D. , Parsa Akbari, Ph.D. , Priyanka Nakka, Ph.D. , Sahar Gelfman, Ph.D. , Sujit Gokhale, B.E. , Tanima De, Ph.D. , Veera Rajagopal, Ph.D. , Alan Shuldiner, M.D. , Bin Ye, Ph.D. , Gannie Tzoneva, Ph.D. , Juan Rodriguez-Flores, Ph.D.

**Research Program Management & Strategic Initiatives**

Esteban Chen, M.S. , Marcus B. Jones, Ph.D. , Michelle G. LeBlanc, Ph.D. , Jason Mighty, Ph.D., Lyndon J. Mitnaul, Ph.D. , Nirupama Nishtala, Ph.D. , Nadia Rana, Ph.D. , Jaimee Hernandez

**Supplemental Methods**

**Protein Measurements**
Protein selection was based on the ability to purify protein targets and published information that the selected proteins are involved in disease pathophysiology. The assay has been described in detail in earlier studies *(16–18)*, and a technical white paper is available from SomaLogic (http://somalogic.com/wp-content/uploads/2017/06/SSM-002-Technical-White-Paper_010916_LSM1.pdf).. The undepleted EDTA-plasma was diluted to 0.05%, 1% and 40% bins.

The samples were then incubated with bin-specific collections of bead-coupled SOMAmers (Slow Off-rate Modified Aptamer reagents that target specific protein targets) in the 96-well-plate format for the washing steps. The washing steps biotinylated the targets and later photocleaved off the beads. The cleaved-off fluorescence-labelled SOMAmers were quantified as a proxy for protein concentration, using custom arrays of SOMAmer-complementary oligonucleotides. Standard samples on each 96-well plate were used to normalize samples on the same and different plates. The normalization process was performed for raw hybridization intensities and median signals, and to bridge between different plates by calibrating the signals.

**Genotype Quality Control and Imputation**

In particular, 400 subjects of African American descent using the Infinium Global Screening Array v2.0. We remove subject with call rate less than 95%, ambiguous gender, excessive and reduced proportion of heterozygosity rate (see figure below) for DNA sample contamination or inbreeding. To further address sample contamination, we examined genome identity-by-descent (IBD), cryptic relationships, and sample duplication. IBD was computed pairwise between all samples using genome-wide genotype data. To avoid genotyping calling error, we excluded SNPs with missing rates > 0.05, minor allele frequency (MAF) < 0.05, and Hardy-Weinberg equilibrium (HWE, p < 1.0 E-03).

After quality control, we excluded 11 subjects and have 386 subjects for the analysis. Quality control steps were carried out using PLINK 1.90b4. Subsequently, we checked the following against the 1000 genome phase 3 (based on the AFR population as reference). Items checked including the incorrect REF/ALT designations, incorrect strand designations, extreme deviations from expected allele frequencies, and palindromic (A/T and G/C) SNPs with allele frequency near 0.5 that are often the source of imputation errors, and generates commands to make files that have fixed or removed these problematic variants. We further used the Michigan imputation server to perform the imputation. The genotypes of the 386 samples were phased with Eagle v2.3, reference panel used was 1000G Phase3 v5. Phase 3 analysis was mapped to GRCh37

**Transfection and Gene Expression**

AC16 cells were transfected with indicated plasmids using LipofectAMINE 2000 (Thermo), according to the manufacturer’s specifications. After 48 hours, cells were either replated for Seahorse assay or collected in TRIzol (Thermo) for RNA isolation. 1ug RNA was reversed transcribed into cDNA using High-Capacity cDNA Reverse Transcription Kit (Thermo Fisher Scientific). The cDNA was used for qRT-PCR analysis using PowerUp SYBR Green master mix (Life Technologies) in a Quant Studio 7 flex (Applied Biosystems). The following primer sequences were used:

FOG2 F TGCTGGACTATCACGAGTGC

FOG2 R GACATCAGGGCTGTTTCGTT

VEGFa F AGGGCAGAATCATCACGAAGT

VEGFa R AGGGTCTCGATTGGATGGCA

ADM F ATGAAGCTGGTTTCCGTCG

ADM R GACATCCGCAGTTCCCTCTT

BNIP3 F CAGGGCTCCTGGGTAGAACT

BNIP3 R CTACTCCGTCCAGACTCATGC

ANGPT1 F CTCGCTGCCATTCTGACTCAC

ANGPT1 R GACAGTTGCCATCGTGTTCTG

ANGPT2 F AATGCAGTACAGAACCAGACG

ANGPT2 R TTAACTTCCGCGTTTGCTCAG

TBP F GAGCCAAGAGTGAAGAACAGTC

TBP R GCTCCCCACCATATTCTGAATCT

36B4 F AGATGCAGCAGATCCGCAT

36B4 R GTTCTTGCCCATCAGCACC

**Tube Formation Assay**

Conditioned media was collected from transfected cells above, spun at 6000g to remove cells and debris and filtered. 10,000 Human Umbilical Vein Endothelial Cells (HUVECs) were seeded onto 60 uL of Growth Factor Reduced Matrigel #354230 (Corning, NY), treated with conditioned media for 6 hours, and imaged under a light microscope at 4x magnification. Images were analyzed using ImageJ Angiogenesis Analyzer (27).

**Oxygen Consumption Rate**

Transfected AC16 cardiomyocytes (30000 cells/well) were treated with 100 uM DMOG for 16 h. These cells were analyzed for mitochondrial metabolism using Seahorse Bioanalyzer Mito Stress Test (Agilent, Wilmington, DE). In brief, Oxygen Consumption rate (OCR) was measured using sequential addition of Oligomycin (1.5 µM), FCCP (1 µM) and Antimycin A/Rotenone mix (0.5 µM). Extracellular Acidification Rate was also determined in parallel to evaluate glycolysis rate. Cells were counted using Hoechst33342 (2 µM) stain on BioTek Cytation 5 Cell Imaging Multimode Reader and used to normalize the OCR and ECAR between groups. Different aspects of metabolism (basal respiration, ATP production, maximal respiration, and spare respiratory capacity) were assessed from OCR as per the manufacturer’s instruction manual and compared between groups.

**Generation of FOG2 knockdown cell line**

FOG2 knockdown lines were generated by genome editing using the CRISPR/Cas9 system as described by Ran et al (28). Briefly, a guide sequence GCTTTGGTGTACAACTACGA was designed using online CRISPR Design Tool (<http://tools.genome-engineering.org>) and cloned into a Cas9-GFP construct. AC16 cells were seeded into a 6 well plate and transfected the next day with 2ug of sgRNA-Cas9-GFP using Lipofectamine 3000 according to manufacturer’s instructions. After 48 hours, the cells were sorted using a Sony SH800s. GFP positive cells were singly plated in a 96 well plate and the individual clones were grown up for screening by PCR for editing and qRT-PCR for FOG2 expression.

**CUT&RUN and DNA Sequencing**

FOG2 knockdown and control lines were incubated under hypoxic conditions overnight (5% O2). CUT&RUN assay kit (CST #86652S) was performed using the following modifications: 144,000 cells were used for each condition and repeated in triplicate. Positive control (Tri-Methyl-Histone, CST#9751), negative control (IgG, CST#66362), and HIF1a (Abcam #2195) were incubated with the samples overnight at 4 degC. Libraries were prepared using the SimpleChip Chip-Seq DNA library prep kit (CST#56795S) and SimpleChIP-seq Multiplex oligos for Illumina (CST#47538S). Anneal and extansion steps were extended to 15 sec x 13 cycles of PCR. Samples were purified using AMPure 1.1X beads (Beckman Coulter) and sequenced on the Illumina NovaSeq 600 using paired-end reads at a depth of 5M.
